# Exploring the Shared Genetic Architectures between Primary Open-Angle Glaucoma and Visual Pathway Regions in the Brain

**DOI:** 10.1101/2025.06.05.25329020

**Authors:** Asma M. Aman, Stuart MacGregor, Santiago Diaz Torres, Puya Gharahkhani

## Abstract

**Purpose:** To investigate the genetic relationships between primary open-angle glaucoma (POAG) and major visual pathways in the brain to better understand the neurological biology of glaucoma, which may facilitate the discovery of neuroprotective drug targets.

**Methods:** We assessed the relationship between POAG and the volumes of five visual pathway regions using genetic correlation and polygenic risk score (PRS). We further used Mendelian randomisation (MR) to investigate the causal relationships. In addition, we used GWAS-pairwise to identify genomic segments with shared causal variants, and summary-data-based MR (SMR) to uncover potential causal genes shared between POAG and the visual brain regions.

**Results:** We found a weak but significant genetic correlation only between POAG and optic chiasm (OC) volume (rg= −0.094, p-value= 0.009). Similarly, PRS for none of the volumes of visual pathway regions showed significant association with POAG, except for the OC volume (OR= 0.96, p-value= 0.0003). In addition, no causal relationships were found between POAG and visual brain regions. The GWAS-pairwise analyses revealed several genomic segments with shared causal variants between POAG and the five brain regions; genetic loci implicated for OC included *CDKN2* gene family region. In addition, the SMR analyses identified five shared potential causal genes, including *PHETA1*, *MAPKAPK5-AS1*, and *EEF1AKMT2*.

**Conclusion:** Our findings suggest a genetic overlap between POAG and the volumes of visual pathway regions, including shared candidate causal genes. Further investigations are required to elucidate the specific role of the shared genes in the aetiology of POAG, and to assess their potential as neuroprotective drug targets.

## Introduction

Primary open-angle glaucoma (POAG) is a progressive and highly heritable optic neuropathy that is a major cause of irreversible vision loss worldwide ^1^. It is characterised by the death of the retinal ganglion cells, which occurs predominantly due to elevated intraocular pressure (IOP) ^2^. Previous research suggested that glaucoma may share common pathways with other neurodegenerative disorders such as Alzheimer’s disease ^3–5^, indicating that POAG aetiology could involve central nervous system pathways. Furthermore, genes related to optic nerve vulnerability are consistently observed in POAG analyses (e.g., *SIX6* and *CDKN2B-AS1*) ^6^, and their effects tend to be more pronounced in normal tension glaucoma ^7^, where IOP remains within the statistically normal range (IOP < 21 mmHg). Research that underlies the aetiology of POAG is mainly focused on IOP, while other mechanisms are largely unexplored. Consequently, there are no current neuroprotective treatments for POAG, and all treatments rely on controlling the IOP to slow down the disease progression; however, they do not prevent the optic neuropathic process.

Observational studies have shown a relationship between POAG and some vision-related brain regions. For instance, studies have demonstrated that the function and structure of the visual cortex, including primary and secondary regions, are affected in glaucoma patients ^8–10^. In addition, previous research has shown that the size of the optic chiasm and lateral geniculate nucleus is smaller in glaucoma patients ^11–14^.

However, there are no comprehensive studies that investigate the genetic overlap and causal relationships between POAG and visual brain regions, particularly in relation to common biological pathways. Therefore, our goal was to investigate the genetic and biological relationships between POAG and visual pathway regions from retina to the visual cortex, as illustrated in **Figure 1**, to better understand the pathophysiology of glaucoma, which may facilitate the discovery of neuroprotective drug targets.

**Figure 1.**
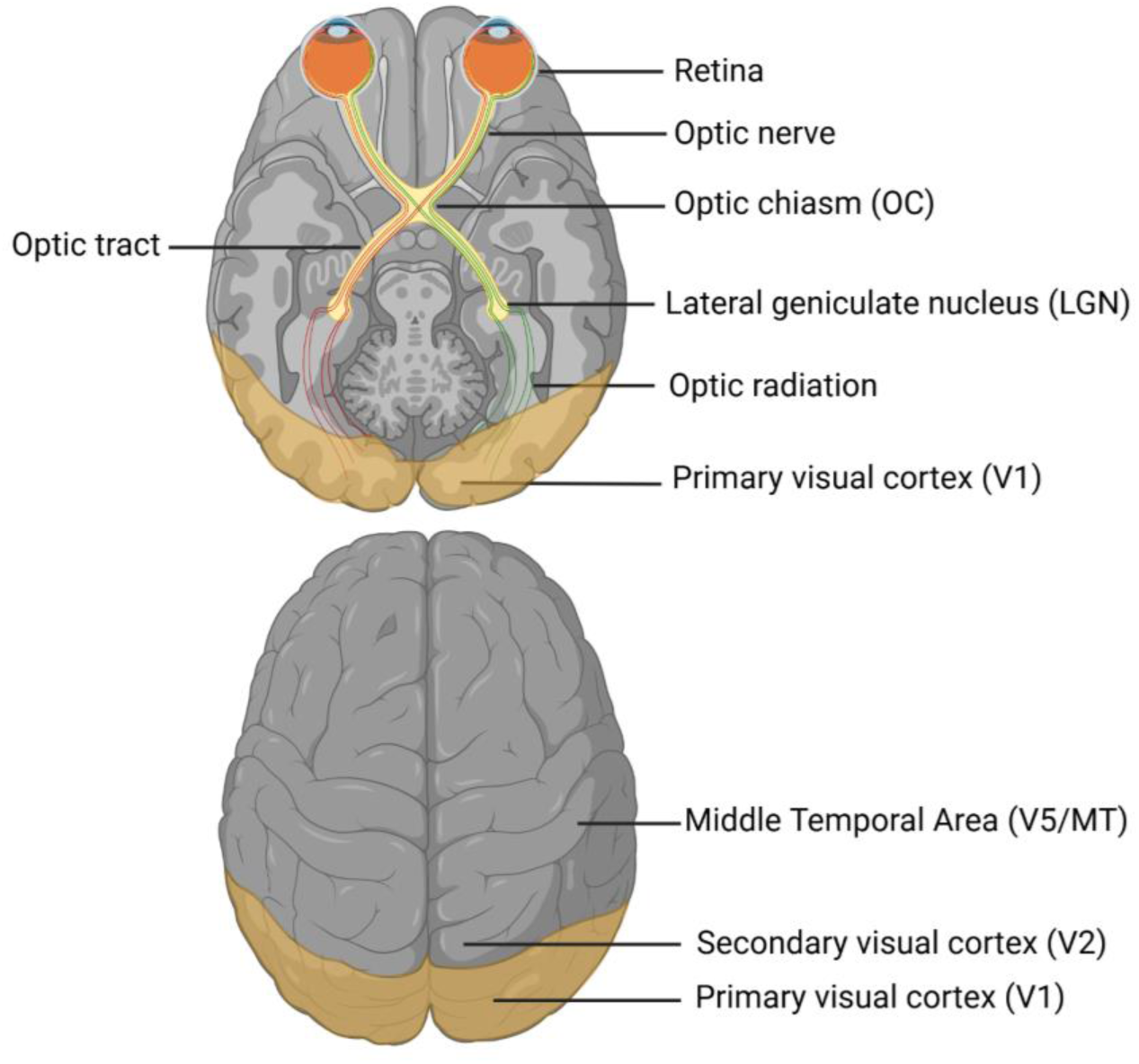
Visual pathway regions, showing the key brain structures involved in the transmission of visual information from the retina to the cortex. That includes the retina, optic nerve, optic chiasm, lateral geniculate nucleus, primary and secondary visual cortex. Created in BioRender. Diaz, S. (2025) https://BioRender.com/gdeng13

## Methods

### Data sources

POAG summary statistics were obtained from a multi-trait genome-wide association study combining genetic information from POAG and two key associated phenotypes, vertical cup-to-disc ratio and IOP, with 644,750 participants, all of European ancestry ^6^.

Genome-wide association studies (GWASs) of visual pathway regions were conducted using around 41,258 European participants from the UK Biobank ^15^. The phenotypes were the volume of optic chiasm (OC) and the mean volume from both the right and left hemispheres for lateral geniculate nucleus (LGN), primary (V1) and secondary visual cortex (V2 and V5). GWASs were conducted using linear mixed models to account for relatedness, with volumes transformed using the rank-based inverse normal transformation method to meet the normality assumption of the models. In addition, the GWASs were adjusted for age, sex, 10 principal components, and intracranial volume. Single nucleotide polymorphisms (SNPs) with minor allele frequency below 0.001 and imputation quality score less than 0.3 were filtered. The analyses were performed using REGENIE software (version 2.2.4).

### Genetic correlation

We used the linkage disequilibrium score regression (LDSC) method to estimate the genetic correlation between POAG and the volume of each visual pathway region. The LDSC is a tool used in statistical genetics which utilises GWAS summary statistics instead of individual-level data and distinguishes between polygenicity and confounding ^16,17^. We used around 5,000 healthy European participants from the UK Biobank as an LD reference. The significance threshold was corrected for multiple testing using the Bonferroni method (p < 0.05/5= 0.01, where five represents the number of visual pathway regions tested).

### Polygenic risk score (PRS)

PRS is a statistical method used to summarise the effects of genetic variants on complex traits. The SBayesRC method ^18^ was used to select credible SNP sets and optimise weights from the GWAS summary statistics, then PLINK (version 2.0) was used to compute the scores. We used the GWAS summary statistics of the volume of each visual pathway region to construct PRS to test for association between these brain regions and the risk of developing POAG. To minimise bias, PRSs were calculated in a subset of individuals who did not overlap with the training set (i.e., those who underwent brain imaging). In addition, first-degree relatives (i.e., kinship coefficient ≥ 0.125) were excluded from the PRS analysis to reduce potential confounding due to relatedness. Logistic regression models were used to assess the associations, adjusting for age and sex. Similarly, we tested for associations between POAG PRS and the volume of each visual pathway region. Here, the POAG PRS was constructed using summary statistics from The Million Veteran Program ^19^, to avoid bias caused by sample overlap. In addition, the volumes were rank-based inverse normal transformed to approximate normality and facilitate comparison between traits. We fitted linear regression models to assess the associations, adjusting for age and sex as covariates. The p-value significance threshold was adjusted using the Bonferroni method (p < 0.005), based on the number of visual pathway regions tested (n=10).

### Causal inference

We explored the putative causality between POAG and the volume of each visual pathway region in both directions using Mendelian randomisation (MR) method. Clumping was performed to select independent genetic instruments for the exposure, using a significance threshold of 5×10⁻⁸ and LD r^2^ threshold of 0.001 to ensure independence between SNPs. We calculated the phenotypic variance explained by the instrumental variables included in the MR analysis for each phenotype, and the variance was ≥ 1%. In cases of significant heterogeneity (i.e., Cochran’s Q test p-value < 0.05), we calculated SNP-level Q statistics (χ², df = 1) and excluded SNPs with high heterogeneity from the MR analysis ^20^. We first applied the inverse variance weighted method ^21^, followed by sensitivity analyses using MR methods, including: MR-Egger regression ^22^, weighted median ^23^, and both simple and weighted mode-based estimators ^24^. Statistical significance was determined to be p < 0.05/10, based on the number of MR tests conducted.

### Colocalisation

It is possible that local genetic overlaps can exist in the absence of genome-wide genetic correlation. We used the GWAS-pairwise (GWAS-PW) approach to identify loci shared between POAG and visual pathway regions ^25^. In this approach, the genome was divided to 1,703 regions and the posterior probability of association (PPA) was computed for each region against four hypotheses: 1) the region contains causal genetic variant exclusively for POAG, 2) the region contains causal genetic variant exclusively for the visual pathway region, 3) the shared region contains common causal genetic variant for POAG and the visual pathway region, and 4) the shared region contains different causal genetic variants for POAG and the visual pathway region. The PPA threshold 0.8 for the third model was used to select the significant shared regions.

### Identifying overlapping causal genes

The multi-SNP summary-data-based Mendelian randomisation (SMR) method was used to test the causality between gene expression and the GWAS analysis for each phenotype ^26,27^, where we used the European UK Biobank reference LD panel. Blood expression quantitative trait loci (eQTL) data were used as genetic instruments for gene expression obtained from eQTLGen Consortium ^28^, which is the largest eQTL database to date with 31,684 participants. In addition, we used eQTL obtained from postmortem retinal tissues ^29^; however, this dataset has limited power (n=453).

We assessed genes located in the shared genomic regions identified by GWAS-PW, adjusting significance threshold for multiple testing using the Bonferroni method (p < 0.05/number of genes in the regions identified by GWAS-PW). The HEIDI test was then used to distinguish between pleiotropy and linkage ^26^, where genes with HEIDI p-value more than 0.05 were considered pleiotropic genes. Afterwards, we extracted and reported the overlapping significant genes between POAG and the visual pathway regions. These genes were then used to identify potential drug repurposing candidates through the Open Targets platform (https://platform.opentargets.org/).

## Results

### Genetic correlation

A weak but significant genetic correlation was found between POAG and OC volume (rg= - 0.094, p-value= 0.009), indicating a slight inverse genetic relationship. However, there was no other significant genetic correlation between POAG and the other visual pathway regions. Genetic correlation results are shown in **Table 1**.

**Table 1.**
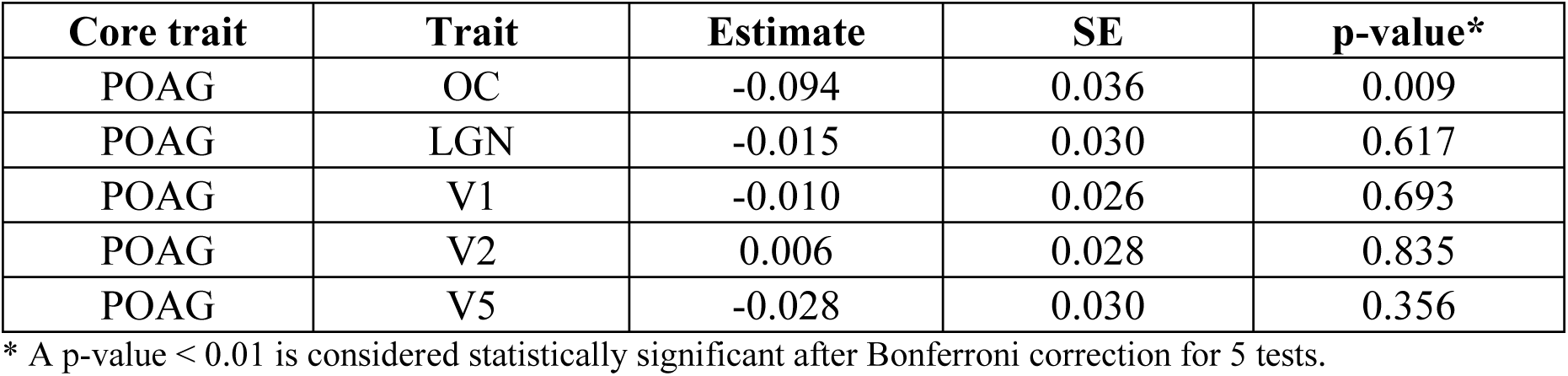
LDSC results for genetic correlations between POAG and volumes of visual pathway regions. Regions analysed include optic chiasm (OC), lateral geniculate nucleus (LGN), primary visual cortex (V1), and secondary visual cortex (V2 and V5).

| Core trait | Trait | Estimate | SE | p-value* |
| --- | --- | --- | --- | --- |
| POAG | OC | -0.094 | 0.036 | 0.009 |
| POAG | LGN | -0.015 | 0.030 | 0.617 |
| POAG | V1 | -0.010 | 0.026 | 0.693 |
| POAG | V2 | 0.006 | 0.028 | 0.835 |
| POAG | V5 | -0.028 | 0.030 | 0.356 |

\* A p-value < 0.01 is considered statistically significant after Bonferroni correction for 5 tests.

### PRS

All PRSs showed no statistically significant result except for the OC volume (OR= 0.96, p-value= 3×10^−4^); complete logistic regression results for PRSs of visual brain regions adjusted for age and sex are presented in **Table 2**. In addition, linear regression results for POAG PRS are provided in the **Supplementary Table 1**. Although the OC PRS was significantly associated with POAG, it did not provide additional predictive value compared to the age and sex model alone (AUC for age and sex model: 0.701, AUC for PRS with age and sex model: 0.702).

**Table 2.** Association between polygenic risk scores for the volumes of visual pathway regions to predict the risk of developing POAG. Regions analysed include optic chiasm (OC), lateral geniculate nucleus (LGN), primary visual cortex (V1), and secondary visual cortex (V2 and V5).

| Trait | Estimate | OR | SE | CI lower | CI upper | p-value* |
| --- | --- | --- | --- | --- | --- | --- |
| OC | -0.036 | 0.964 | 0.010 | 0.946 | 0.984 | $3 \times 10^{-4}$ |
| LGN | -0.017 | 0.983 | 0.010 | 0.964 | 1.003 | 0.093 |
| V1 | 0.017 | 1.017 | 0.010 | 0.997 | 1.038 | 0.085 |
| V2 | 0.009 | 1.009 | 0.010 | 0.990 | 1.029 | 0.352 |
| V5 | 0.008 | 1.008 | 0.010 | 0.988 | 1.028 | 0.423 |

\* A p-value < 0.005 is considered statistically significant after Bonferroni correction for 10 tests.

### Causal inference

The MR analyses revealed no statistically significant causal association between POAG and the volumes of visual pathway regions in either direction. MR results using the inverse variance weighted method are presented in **Tables 3 and 4**, while MR sensitivity analysis results and plots are available in the **Supplementary Tables 2-11 and Supplementary Figures 1-2**, respectively.

**Table 3.** Mendelian randomisation results using the inverse variance weighted method to test causality between POAG as an outcome and volumes of visual pathway regions as exposures. Regions analysed include optic chiasm (OC), lateral geniculate nucleus (LGN), primary visual cortex (V1), and secondary visual cortex (V2 and V5).

| Exposure | Outcome | Beta | SE | p-value* |
| --- | --- | --- | --- | --- |
| OC | POAG | -0.077 | 0.093 | 0.409 |
| LGN | POAG | -0.055 | 0.100 | 0.578 |
| V1 | POAG | 0.001 | 0.033 | 0.981 |
| V2 | POAG | -0.063 | 0.041 | 0.123 |
| V5 | POAG | 0.081 | 0.063 | 0.201 |

\* A p-value < 0.005 is considered statistically significant after Bonferroni correction for 10 tests.

**Table 4.** Mendelian randomisation results using the inverse variance weighted method to test causality between POAG as an exposure and volumes of visual pathway regions as outcomes. Regions analysed include optic chiasm (OC), lateral geniculate nucleus (LGN), primary visual cortex (V1), and secondary visual cortex (V2 and V5).

| Exposure | Outcome | Beta | SE | p-value* |
| --- | --- | --- | --- | --- |
| POAG | OC | -0.013 | 0.007 | 0.063 |
| POAG | LGN | -0.014 | 0.006 | 0.015 |
| POAG | V1 | -0.002 | 0.006 | 0.783 |
| POAG | V2 | -0.009 | 0.006 | 0.136 |
| POAG | V5 | -0.003 | 0.006 | 0.592 |

\* A p-value < 0.005 is considered statistically significant after Bonferroni correction for 10 tests.

### Colocalisation

The GWAS-PW analysis identified eight shared genomic regions with causal variants common to both POAG and OC, where the region on chromosome 9 containing *CDKN2* gene family is among the identified regions. This gene family has an established association with POAG in previous studies ^6^. In addition, six regions were shared with V2, five regions with V1, and one region each with V5 and LGN. The details of the significant shared regions are available in the **Supplementary Table 12**.

### SMR

The SMR analyses, using eQTLGen, revealed five potential causal genes shared between POAG and visual pathway regions that met the significance criteria (i.e., SMR p-value < 0.05/ number of genes in the GWAS-PW identified regions, and HEIDI test p > 0.05) and located within regions highlighted by GWAS-PW. In particular, the analyses identified a few significant genes shared with V1 including *PHETA1*, a gene involved in endosomal trafficking and may play a role in regulating membrane dynamics by interacting with inositol polyphosphate 5-phosphatase *OCRL-1* enzyme ^30^, and *MAPKAPK5-AS1*, a long non-coding RNA (lncRNA) gene that has a role in cancer and rheumatoid arthritis ^31–33^. Furthermore, the *EEF1AKMT2* gene was found to be shared between POAG and V2, which is involved in protein methylation ^34,35^. **Table 5** presents the full list of the shared potential causal genes. The identified potential causal genes were not targets of existing drugs in the Open Targets platform. Also, the SMR analyses using retinal tissues have not identified any significant results.

**Table 5.** Significant genes shared between POAG and visual pathway regions identified by SMR analysis (SMR p-value survived multiple testing and HEIDI test p > 0.05) and located within GWAS-PW highlighted regions.

| Trait | Gene | Location | Trait |  |  | POAG |  |  |
| --- | --- | --- | --- | --- | --- | --- | --- | --- |
|  |  |  | SMR estimate (SE) | SMR p-value* | HEIDI p-value | SMR estimate (SE) | SMR p-value* | HEIDI p-value |
| V1 | ENSG00000198324<br>(*PHETA1*,<br>*FAM109A*) | Chr12,<br>chunk<br>1245 | 0.174<br>(0.049) | $2 \times 10^{-4}$ | 0.810 | 0.197<br>(0.069) | $3 \times 10^{-4}$ | 0.337 |
| V1 | ENSG00000234608<br>(*MAPKAPK5-AS1*) | Chr12,<br>chunk<br>1245 | 0.061<br>(0.018) | $2 \times 10^{-7}$ | 0.069 | 0.069<br>(0.025) | $2 \times 10^{-7}$ | 0.061 |
| V1 | ENSG00000204852<br>(*TCTN1*) | Chr12,<br>chunk<br>1245 | -0.020<br>(0.055) | $7 \times 10^{-4}$ | 0.324 | 0.102<br>(0.078) | $6 \times 10^{-6}$ | 0.056 |
| V2 | ENSG00000203791<br>(*EEF1AKMT2*,<br>*METTL10*) | Chr10,<br>chunk<br>1087 | -0.125<br>(0.027) | $6 \times 10^{-5}$ | 0.157 | -0.222<br>(0.043) | $3 \times 10^{-5}$ | 0.586 |
| V2 | ENSG00000257877 | Chr12,<br>chunk<br>1245 | 0.028<br>(0.017) | $2 \times 10^{-4}$ | 0.080 | 0.081<br>(0.028) | $3 \times 10^{-5}$ | 0.280 |

\* A p-value $< 0.05$ / the number of genes present in the regions of interest identified by GWAS-PW is considered statistically significant after Bonferroni correction. In particular, adjustments were conducted for 121 genes in OC, 5 in LGN, 62 in V1, 61 in V2, and 15 in V5.

## Discussion

To date, there are no studies investigating the genetic overlap between POAG and the regions of the visual pathway across the genome. Here, we explored the shared genetic architecture between five brain regions and POAG. In particular, we examined the genetic correlation, PRS, causal association, colocalisation, and shared potential causal genes between POAG and the volumes of those brain areas in the shared genomic regions. The results revealed significant genetic correlation and PRS association between OC and POAG, while the other visual regions did not show significant findings. However, GWAS-PW identified a few shared genetic regions between POAG and the five visual regions, and SMR analyses identified five shared candidate causal genes.

POAG primarily affects the optic nerve, and the current literature hints that it might also affect the visual pathway regions in the brain ^8–14^. In this study, we show that despite the fact that no statistically significant causal association was observed, the effect sizes were consistent in their direction. However, the results remain inconclusive, likely because the power to detect causality for small to modest effect sizes was limited due to small sample size.

The genetic correlation and PRS results indicate that OC is biologically related to POAG. That could be due to the fact that OC is an extension of the optic nerve that is damaged during POAG-related neuropathy. The direction of the association observed in our analyses aligns with the findings from observational studies, which showed that individuals with POAG have decreased OC size, indicating an inverse relationship between both traits ^11–13^.

The segment on chromosome 12, ranging from position 110,336,875 to 113,261,782, includes four potential causal genes. One gene is *PHETA1*, which is associated with cup-to-disc ratio ^36^, a trait indicating optic nerve degeneration and is closely related to POAG. Another gene is *MAPKAPK5-AS1*, which is a lncRNA gene (i.e., lncRNAs are important regulators of gene expression and cellular processes) that has been shown to promote diseases through regulating the adjacent gene *MAPKAPK5* ^37^. *MAPKAPK5* gene has a role in neuronal plasticity and cell cycle ^38^. It has also been associated with Alzheimer’s disease pathogenesis, and it has been presented as a potential biomarker for cognitive decline ^39,40^. Also, both genes were previously associated with POAG ^6^; however, further research in the specific roles of *PHETA1* and *MAPKAPK5-AS1* genes in POAG could facilitate the development of novel therapeutic molecules targeting these genes.

The region on chromosome 10 contains one candidate causal gene. *EEF1AKMT2* gene encodes a protein that functions as protein-lysine N-methyltransferase, an enzyme that regulates gene expression and cellular processes through post-translational modification ^34,35^. The regulation of protein synthesis is vital for all cells’ health and survival, including neurons. This gene was found to be associated with the hippocampus volume, a region implicated in brain diseases such as Alzheimer’s disease and schizophrenia ^41,42^. Given that this gene was also mapped for POAG ^6^, this observation highlights the need for further investigation into the role of *EEF1AKMT2* gene in glaucoma aetiology and as a treatment target.

This study is limited to the European population, which may lack the genetic diversity present in other ethnic groups. As a result, the findings cannot be generalised to other populations. In addition, the power of MR analysis to detect small to modest causal relationships may have been limited by the sample size of POAG and the brain regions. Similarly, the sample size of the volumes of visual pathway regions is relatively small, which potentially limits the statistical power to detect genetic variants. Larger cohorts would provide better power to identify additional overlapping genes and more robust biological pathways. While we identified certain genes associated with POAG and visual pathways, functional validation of these genes was not performed. Without experimental validation, the biological relevance of these associations remains uncertain. Further studies are needed to confirm the functional impact of the identified genes.

Our findings suggest a genetic overlap between POAG and the volumes of visual pathway regions in the brain, including shared candidate causal genes. Further investigations are required to elucidate the specific role of the shared genes in the aetiology of POAG, and to assess their potential as neuroprotective drug targets.

## Data Availability

UK Biobank data are available to qualified researchers upon application to UK Biobank. POAG summary statistics were obtained from publicly available sources.

## Acknowledgments

This research has been conducted using the UK Biobank data under project number 25331. S.M. (2034568) and P.G. (1173390) are supported by Investigator grants from the Australian National Health and Medical Research Council (NHMRC).

## Supplementary Tables

**Supplementary Table 1.** Association between polygenic risk score for POAG to predict the volume of each visual pathway region.

**Supplementary Table 2.** Mendelian randomisation analysis results for optic chiasm on POAG.

**Supplementary Table 3.** Mendelian randomisation analysis results for lateral geniculate nucleus (LGN) on POAG.

**Supplementary Table 4.** Mendelian randomisation analysis results for primary visual cortex (V1) on POAG.

**Supplementary Table 5.** Mendelian randomisation analysis results for secondary visual cortex (V2) on POAG.

**Supplementary Table 6.** Mendelian randomisation analysis results for secondary visual cortex (V5) on POAG.

**Supplementary Table 7.** Mendelian randomisation analysis results for POAG on optic chiasm.

**Supplementary Table 8.** Mendelian randomisation analysis results for POAG on lateral geniculate nucleus (LGN).

**Supplementary Table 9.** Mendelian randomisation analysis results for POAG on primary visual cortex (V1).

**Supplementary Table 10.** Mendelian randomisation analysis results for POAG on secondary visual cortex (V2).

**Supplementary Table 11.** Mendelian randomisation analysis results for POAG on secondary visual cortex (V5).

**Supplementary Table 12.** Significant shared regions between POAG and Visual pathway regions with PPA3 > 0.8.

## Supplementary Figures

**Supplementary Figure 1.** Mendelian Randomization analysis evaluating the causal effect of the volume of each visual pathway region (i.e., exposure) on POAG (i.e., outcome).

**Supplementary Figure 2.** Mendelian Randomization analysis evaluating the causal effect of POAG (i.e., exposure) on the volume of each visual pathway region (i.e., outcome).

