## Supplementary Figures for "Exploring the Shared Genetic Architectures between Primary Open-Angle Glaucoma and Visual Pathway Regions in the Brain"

**Supplementary Figure 1.** Mendelian Randomization analysis evaluating the causal effect of the volume of each visual pathway region (i.e., exposure) on POAG (i.e., outcome). Each point represents a genetic variant as instrumental variable for the exposure, with the X-axis showing the effect on the exposure and the Y-axis showing the effect on the outcome. The lines indicate the estimated causal effect derived from the following methods: inverse variance weighted, MR-Egger, simple mode, both weighted median and mode.

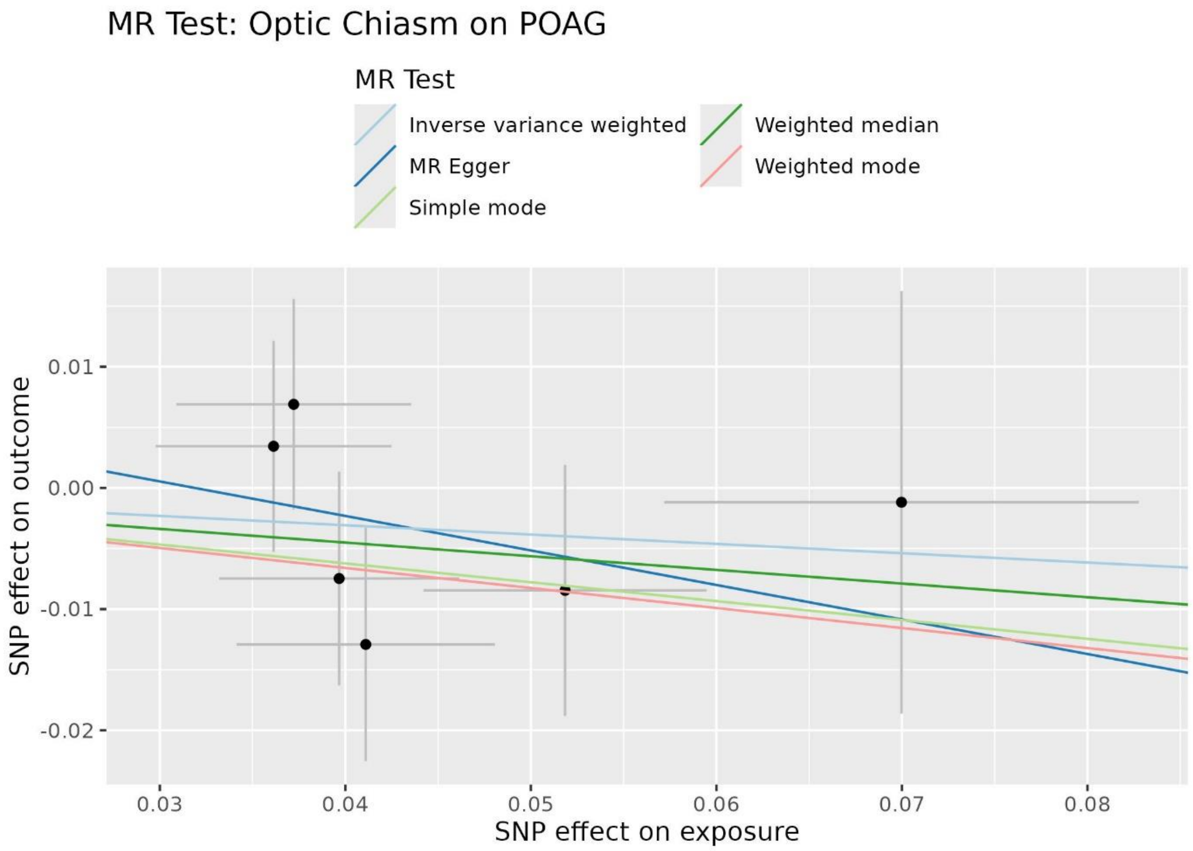

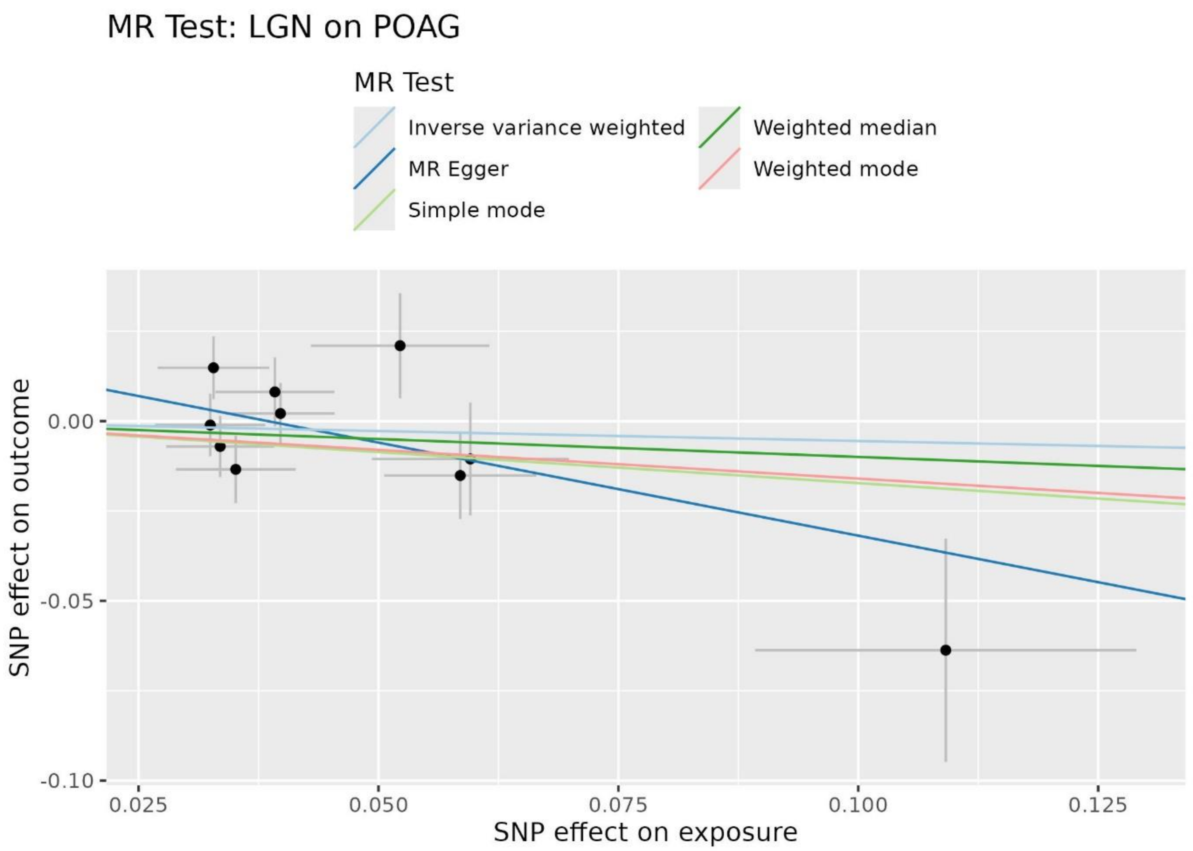
MR Test: LGN on POAG
MR Test
Inverse variance weighted
MR Egger
Simple mode
Weighted median
Weighted mode
SNP effect on outcome
0.00
-0.05
-0.10
0.025
0.050
0.075
0.100
0.125
SNP effect on exposure

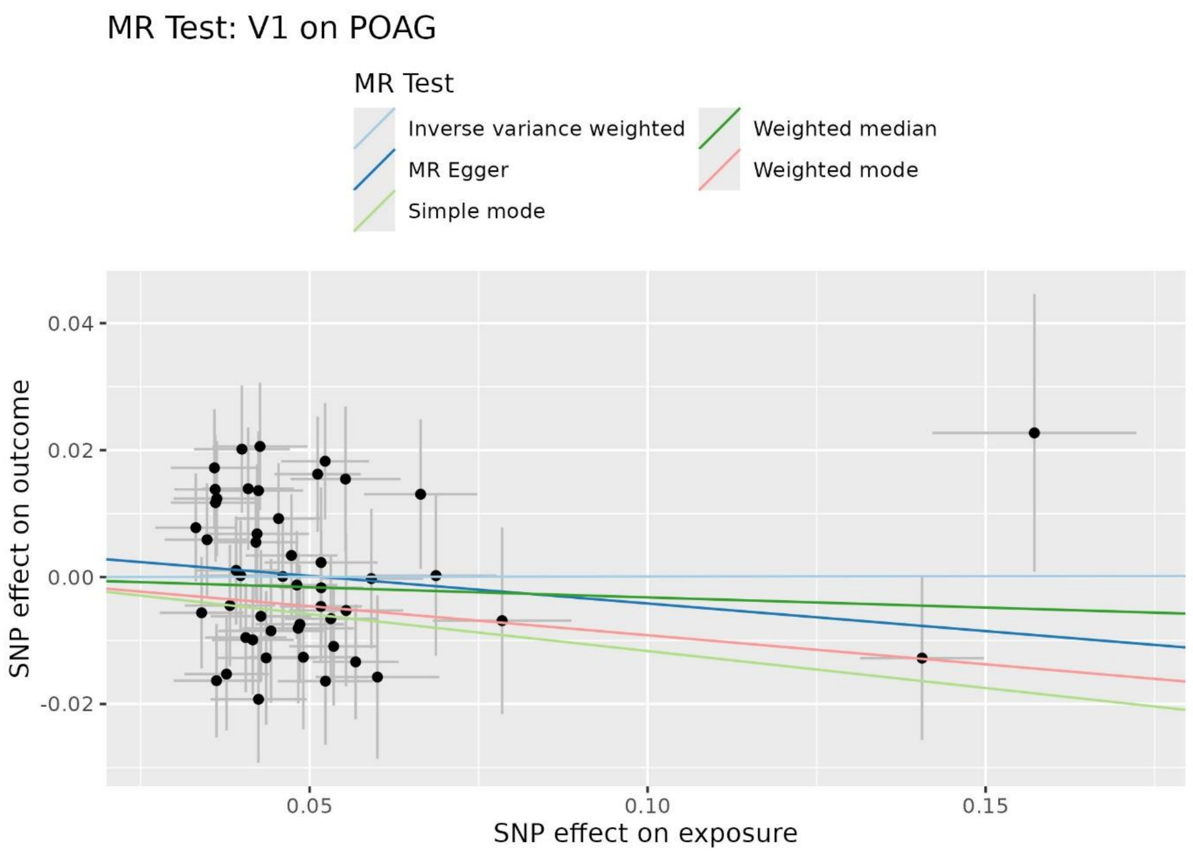
MR Test: V1 on POAG
MR Test
Inverse variance weighted
MR Egger
Simple mode
Weighted median
Weighted mode
SNP effect on outcome
0.04
0.02
0.00
-0.02
0.05
0.10
0.15
SNP effect on exposure

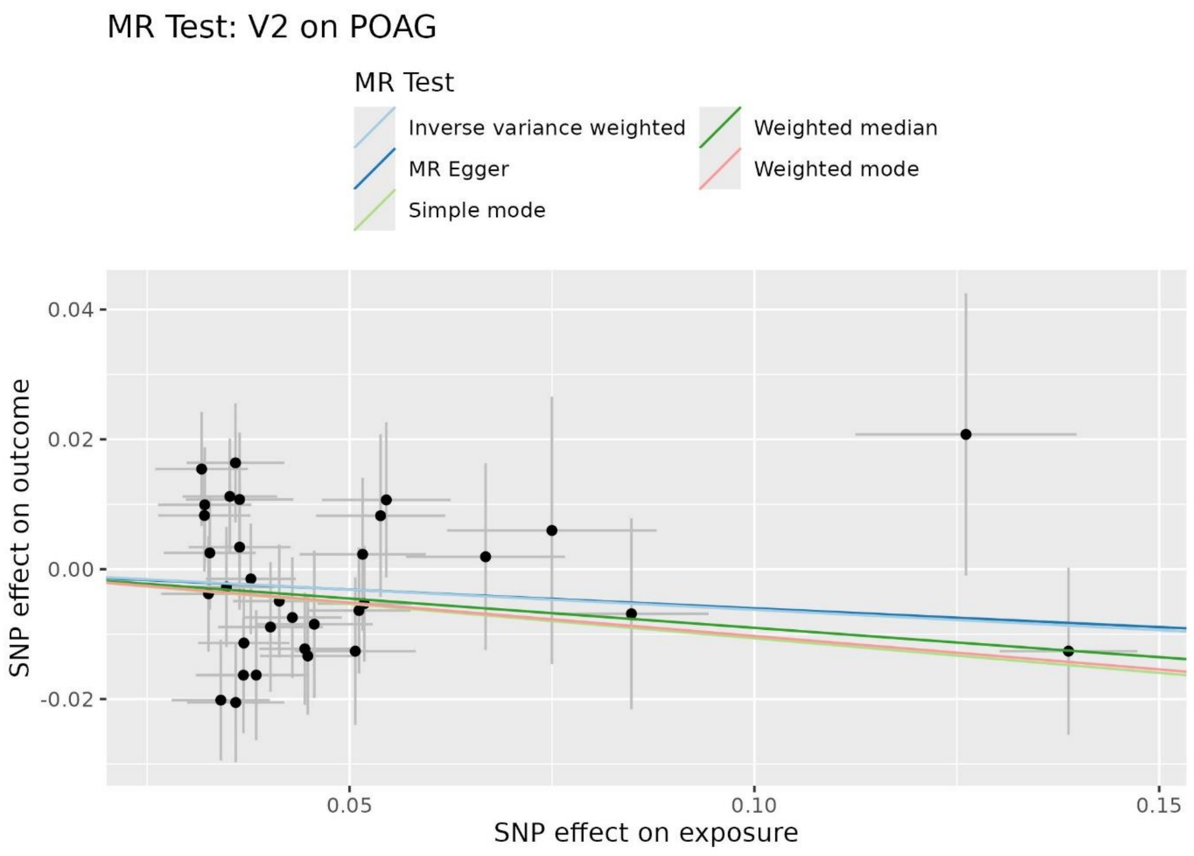
MR Test: V2 on POAG
MR Test
Inverse variance weighted
MR Egger
Simple mode
Weighted median
Weighted mode
0.04
0.02
0.00
-0.02
SNP effect on outcome
0.05
0.10
0.15
SNP effect on exposure

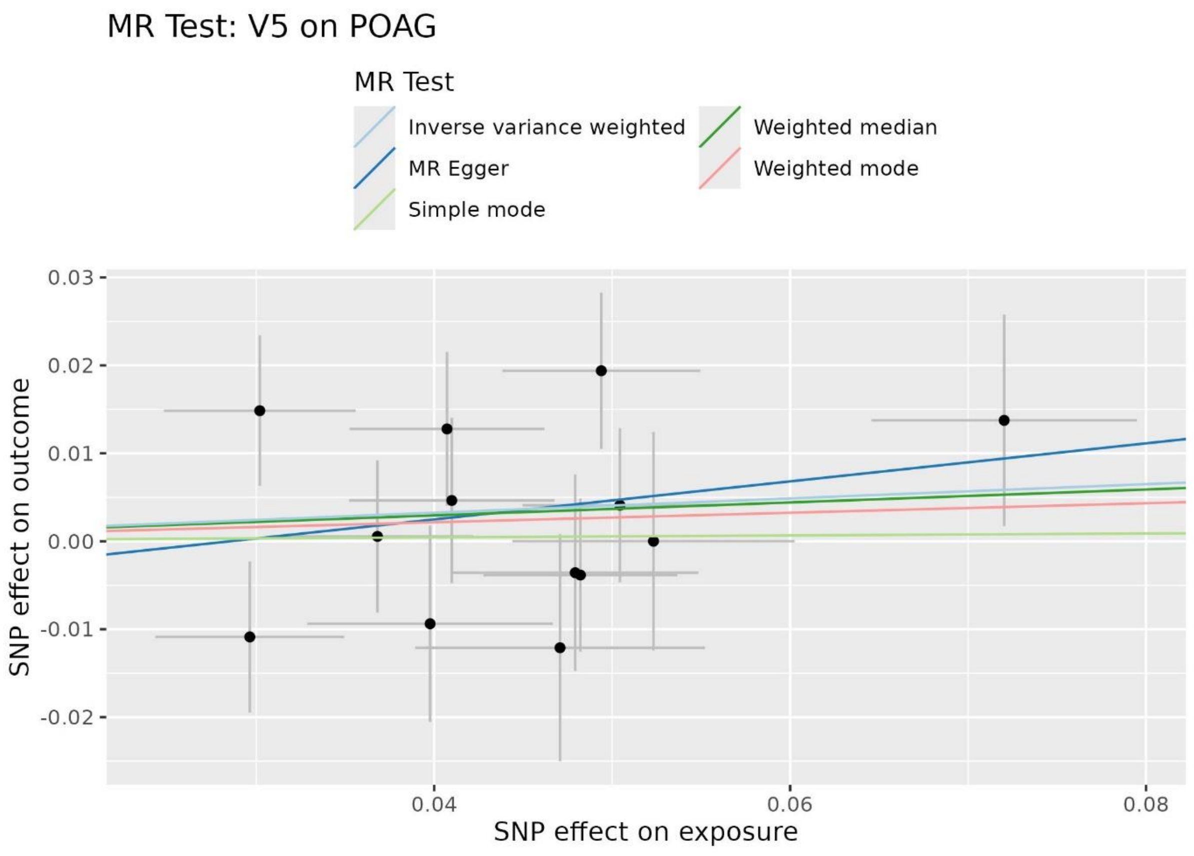
MR Test: V5 on POAG
MR Test
Inverse variance weighted
MR Egger
Simple mode
Weighted median
Weighted mode
0.03
0.02
0.01
0.00
-0.01
-0.02
SNP effect on outcome
0.04
0.06
0.08
SNP effect on exposure

**Supplementary Figure 2.** Mendelian Randomization analysis evaluating the causal effect of POAG (i.e., exposure) on the volume of each visual pathway region (i.e., outcome). Each point represents a genetic variant as instrumental variable for the exposure, with the X-axis showing the effect on the exposure and the Y-axis showing the effect on the outcome. The lines indicate the estimated causal effect derived from the following methods: inverse variance weighted, MR-Egger, simple mode, both weighted median and mode.

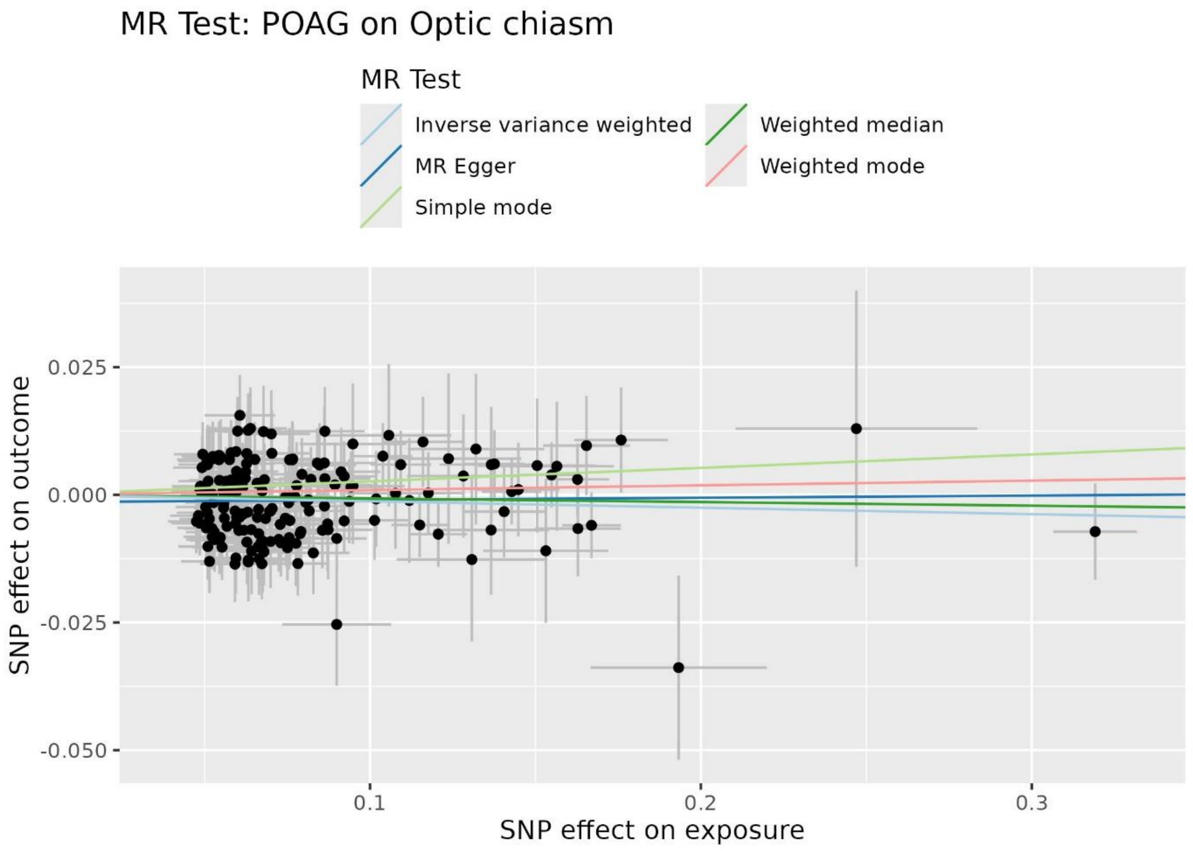

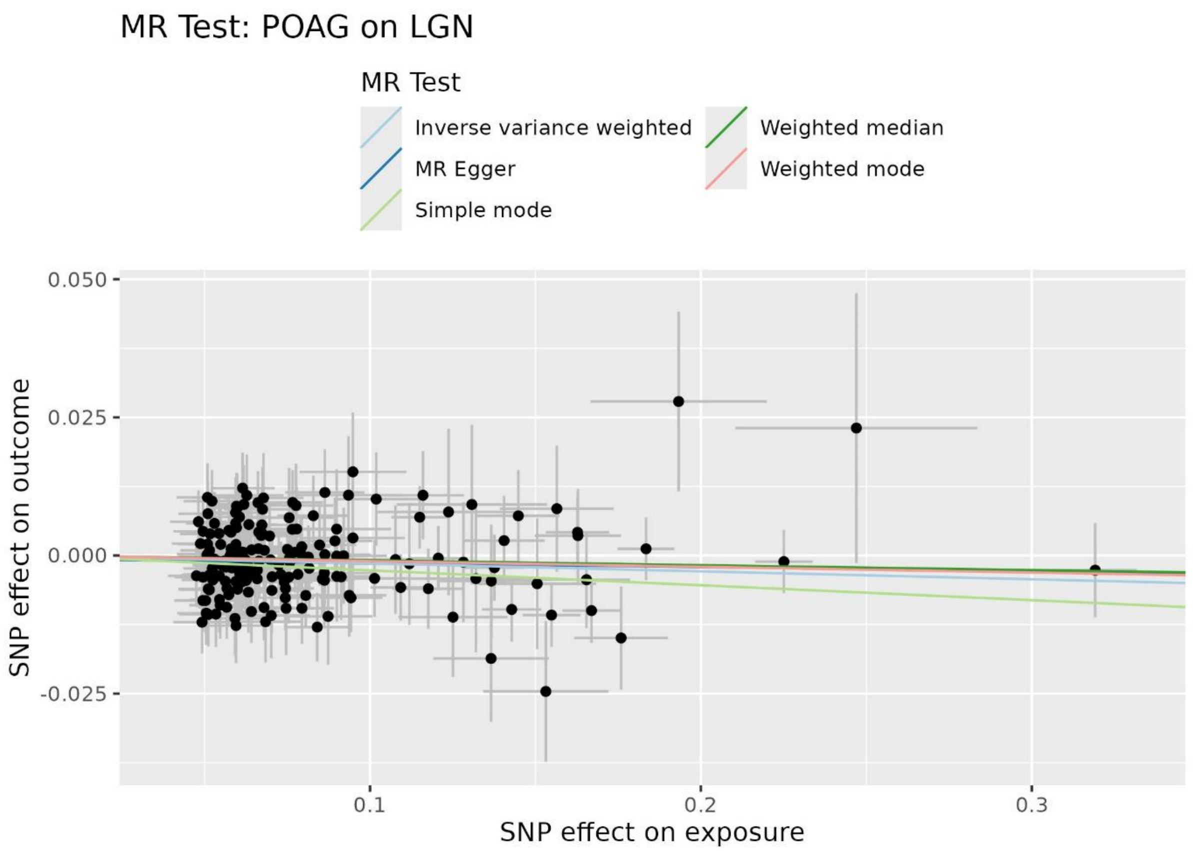
MR Test: POAG on LGN
MR Test
Inverse variance weighted
MR Egger
Simple mode
Weighted median
Weighted mode
0.050
0.025
0.000
-0.025
SNP effect on outcome
0.1
0.2
0.3
SNP effect on exposure

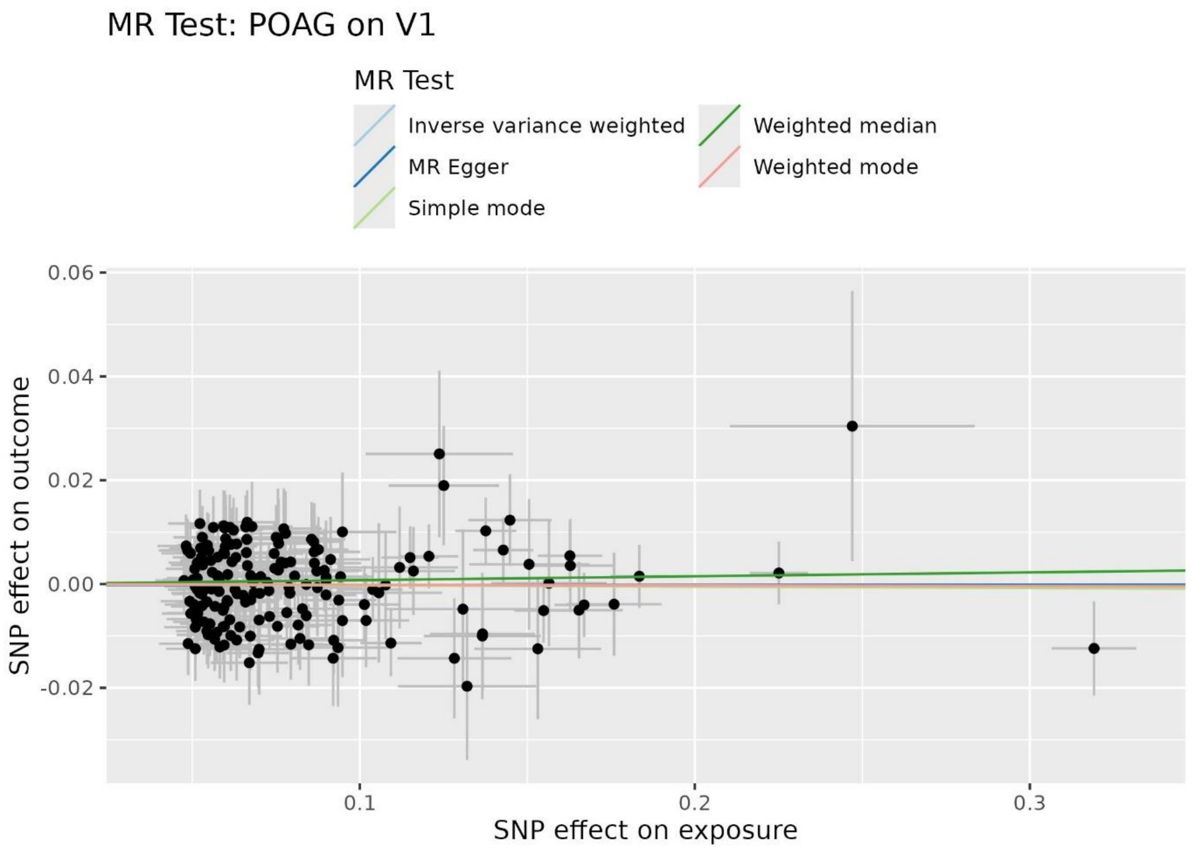
MR Test: POAG on V1
MR Test
Inverse variance weighted
MR Egger
Simple mode
Weighted median
Weighted mode
0.06
0.04
0.02
0.00
-0.02
SNP effect on outcome
0.1
0.2
0.3
SNP effect on exposure

### MR Test: POAG on V2

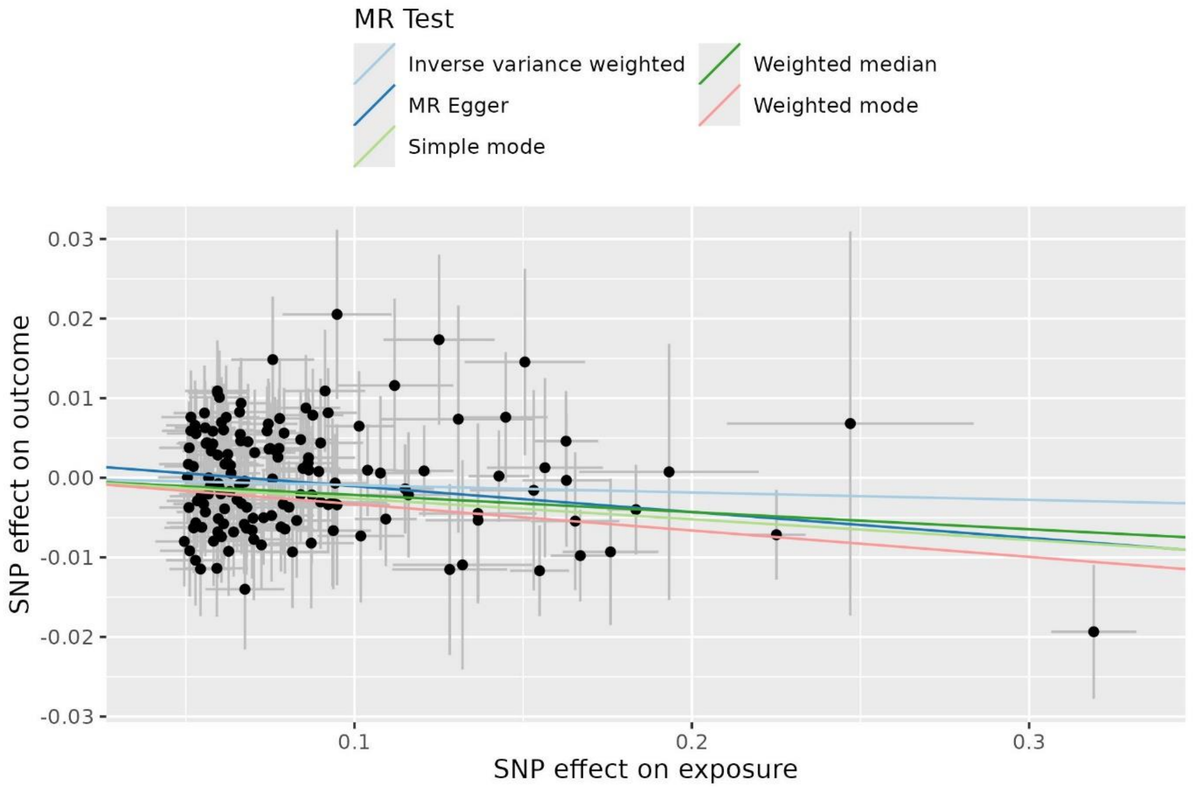


### MR Test: POAG on V5

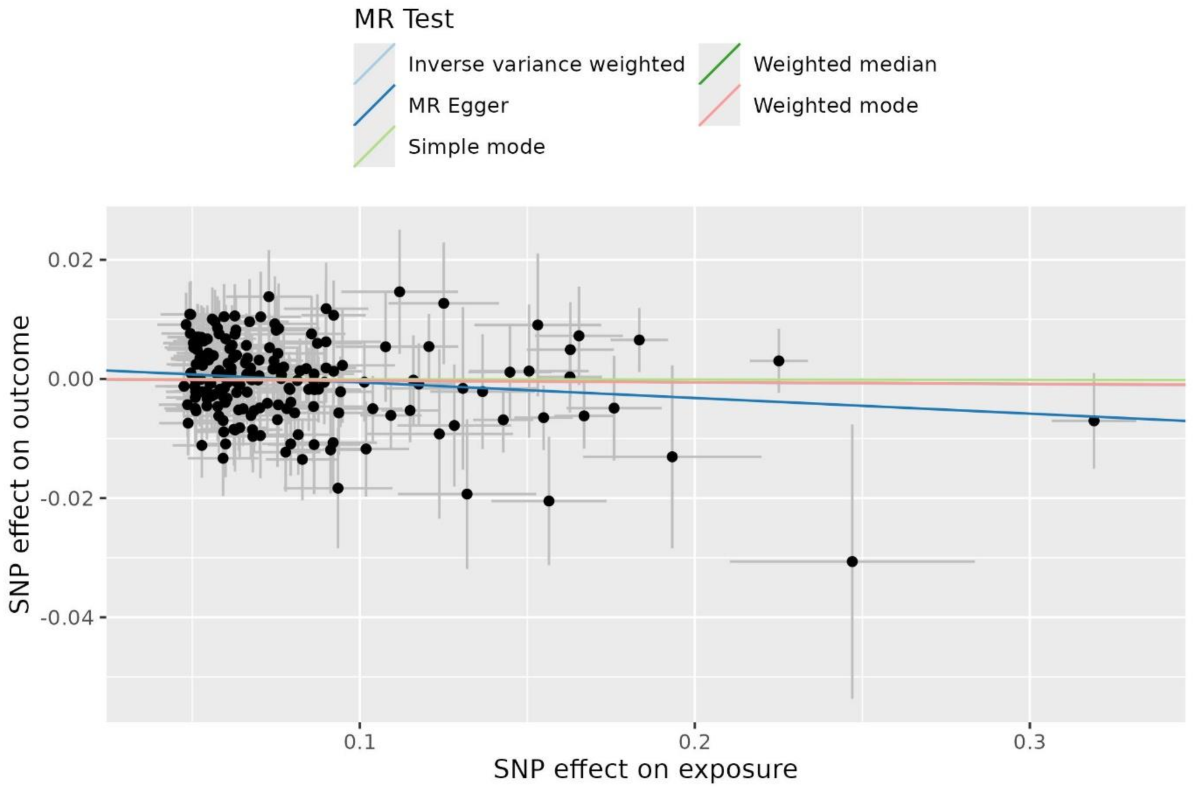
